# Time intervals between COVID-19 cases, and more severe outcomes

**DOI:** 10.1101/2022.10.31.22281769

**Authors:** A.J. Wood, R.R. Kao

**Affiliations:** Roslin Institute, University of Edinburgh; Royal (Dick) School of Veterinary Studies, University of Edinburgh

## Abstract

A critical factor in infectious disease control is the risk of an outbreak overwhelming local healthcare capacity. The overall demand on healthcare services will depend on disease severity, but the precise timing and size of peak demand also depends on the time *interval* (or clinical time delay) between initial infection, and development of severe disease. A broader *distribution* of intervals may draw that demand out over a longer period, but have a lower peak demand. These interval distributions are therefore important in modelling trajectories of e.g. hospital admissions, given a trajectory of incidence. Conversely, as testing rates decline, an incidence trajectory may need to be inferred through the delayed, but relatively unbiased signal of hospital admissions.

Healthcare demand has been extensively modelled during the COVID-19 pandemic, where localised waves of infection have imposed severe stresses on healthcare services. While the initial acute threat posed by this disease has since subsided from immunity buildup from vaccination and prior infection, prevalence remains high and waning immunity may lead to substantial pressures for years to come. In this work, then, we present a set of interval distributions, for COVID-19 cases and subsequent severe outcomes; hospital admission, ICU admission, and death. These may be used to model more realistic scenarios of hospital admissions and occupancy, given a trajectory of infections or cases.

We present a method for obtaining empirical distributions using COVID-19 outcomes data from Scotland between September 2020 and January 2022 (*N* = 31724 hospital admissions, *N* = 3514 ICU admissions, *N* = 8306 mortalities). We present separate distributions for individual age, sex, and deprivation of residing community. We show that, while the risk of severe disease following COVID-19 infection is substantially higher for the elderly or those residing in areas of high deprivation, the length of stay shows no strong dependence, suggesting that severe outcomes are equally severe across risk groups. As Scotland and other countries move into a phase where testing is no longer abundant, these intervals may be of use for retrospective modelling of patterns of infection, given data on severe outcomes.

## 1 Introduction

The threat posed by an infectious disease on a human population depends critically on the risk of developing severe illness, and the burden that may place on healthcare systems. During the early stages of transmission or a new wave of infection, it is important to understand when and how to expect that future demand on healthcare systems to come. Depending on the natural history of a particular disease, this may be immediate and acute, or be drawn out over a longer period of time.

Such estimation of future healthcare burden has been particularly important during the COVID-19 pandemic, where severe curbs on everyday life were imposed globally, in order to control the spread of infection and prevent the overwhelming of hospital capacity. Now over two years from the first known case, the acute threat posed by the virus has subsided, from the build-up of population immunity via effective vaccination and prior infection. However, future pressures on healthcare services due to COVID-19 may still be considerable, with a continuing circulation of infection and waning immunity on timescales of order six months [1].

In prior waves of COVID-19 infection, the peak demand on healthcare services has followed, with a delay, after a peak in incidence. Our work here focuses on the length of this delay and its dependence on age and sex; risk factors that are known to be important for determining the likelihood of severe infection. For modelling future waves of infection, the delay between infection and developing of severe disease (and variation in that delay from person to person) introduces uncertainty on the timing and size of the future *peak* demand on healthcare services, even if an overall infection-hospitalisation rate is well estimated.

The COVID-19 pandemic has been unique in the volume and granularity of data recorded for a human disease. This is not just for hospital admissions and deaths (for which data are already routinely collected for many illnesses), but on proactive testing for the disease. Using such data, the aim of this work is to then obtain empirical distributions of the time intervals between different outcomes at the individual level. We use data collected across Scotland, between September 2020 and January 2022. In this period, COVID-19 tests were widely and freely available, with reported results collected in central databases.

The Scottish COVID-19 data are advantageous for our study as they include additional identifiers in the data, allowing us to show how these distributions differ by a person’s age, sex, and deprivation in their area of residence, of which all are known risk factors for poor COVID-19 outcomes [2, 3, 4, 5, 6, 7]. Our distributions build on length-of-stay distributions obtained from the first year of the pandemic [8, 9, 10, 11, 12, 13, 14], as well as inferred distributions of intervals from the initial infection stage [15], and from onset of symptoms to diagnosis or mortality [16].

The first set of distributions we describe are between COVID-19 cases, and three different severe outcomes: hospital admission, intensive care unit (ICU) admission, and mortality. These affect the shape of the trajectory of severe outcomes, given a trajectory of cases. Interval distributions with a higher mean value will lead to a greater delay between cases and following severe outcomes, whereas a higher variance will lead to a more drawn-out trajectory, with a lower peak.

The second set of distributions are those *between* different severe outcomes. These relate to hospital occupancy; how long an individual spends in hospital once admitted. While the data do not specify recoveries or discharges, we also estimate the distribution of intervals between admission and discharge, for patients admitted that go on to *survive*, using public data on hospital occupancy. We infer that over time, an increasing proportion of COVID-19 hospital burden in the period studied was of individuals that eventually went on to be discharged, and the mean time spent in hospital shortened.

## 2 Data

COVID-19 data are provided by Public Health Scotland’s (PHS) electronic Data Research and Innovation Service (eDRIS) under a data sharing agreement, and specify COVID-19 tests and severe outcomes. Each entry has an associated date, and de-identified patient ID with age (in five-year windows, up to 75+), sex, and residing *data zone* (DZ). DZs are non-overlapping Scottish census areas, each with a residing population of order 500–1,000. There are 6,976 DZs in total, covering the full area and population of Scotland. A subpopulation of a certain DZ, age range, and sex (e.g., men aged between 50–54 living in a particular DZ) will typically have 0–50 individuals, allowing us to analyse outcomes at this same resolution.

The test data contain results from both rapid lateral flow device (LFD) tests and polymerase chain reaction (PCR) tests. In the period studied, public health policy was that those exhibiting COVID-19 symptoms or testing positive on an LFD should report the result, and take a confirmatory PCR test (usually at a local testing centre). We define a case as a new positive test result, taken at least 60 days from any previous positive test by the same individual. For those with repeat positive tests within 60 days (such as a LFD positive followed by a PCR positive), we use the date of the first PCR positive, or the first LFD positive if there is no PCR positive. The case date is the date on which the test is taken.

A *severe* COVID-19 outcome — a hospital admission, intensive care unit (ICU) admission, or mortality — is one where “*coronavirus*” or “*COVID-19* “ appears within the underlying causes for that outcome. This analysis therefore includes severe outcomes “with” COVID-19. The date associated with hospitalisation and ICU admission is the date of first admission, and the date associated with mortality is the day the individual dies. We assume that if an individual dies after a hospitalisation or ICU admission, then they died in hospital, and were in hospital for the whole time.

Outcomes between September 10 2020 and January 6 2022 were considered. In this period of the epidemic testing was both free, and widely available to the whole population in Scotland. We exclude data for those aged under 20, owing to a scarcity of severe outcomes. Presented values draw from the version of the eDRIS data dated October 27 2022.

Data on overall COVID-19 hospital occupancy are obtained from PHS, with daily figures on patients in hospital with confirmed COVID-19, throughout the period studied [17].

In our analysis we define an *interval* Δ*t*_AB_ as the elapsed time between two different outcomes A and B. This is given as a whole number of days. We obtain distributions for:

- *Case* intervals, between cases and more severe outcomes: Case-to-Hospitalisation (Δ*t*_CH_), Case-to-ICU admission (Δ*t*_CI_), Case-to-Mortality (Δ*t*_CM_), and;
- *Nosocomial* intervals, between different outcomes while in hospital, affecting length of stay: Hospitalisation-to-ICU admission (Δ*t*_HI_), Hospitalisation-to-Mortality (Δ*t*_HM_), ICU admission-to-Mortality (Δ*t*_IM_).

We detail in Appendix A how the high resolution of the data allows different outcomes to be associated.

## 3 Results

### 3.1 Interval distributions

Fig. 1 summarises all empirical interval distributions, alongside parameters for distribution fits (detailed in Appendix B).

**Figure 1:**
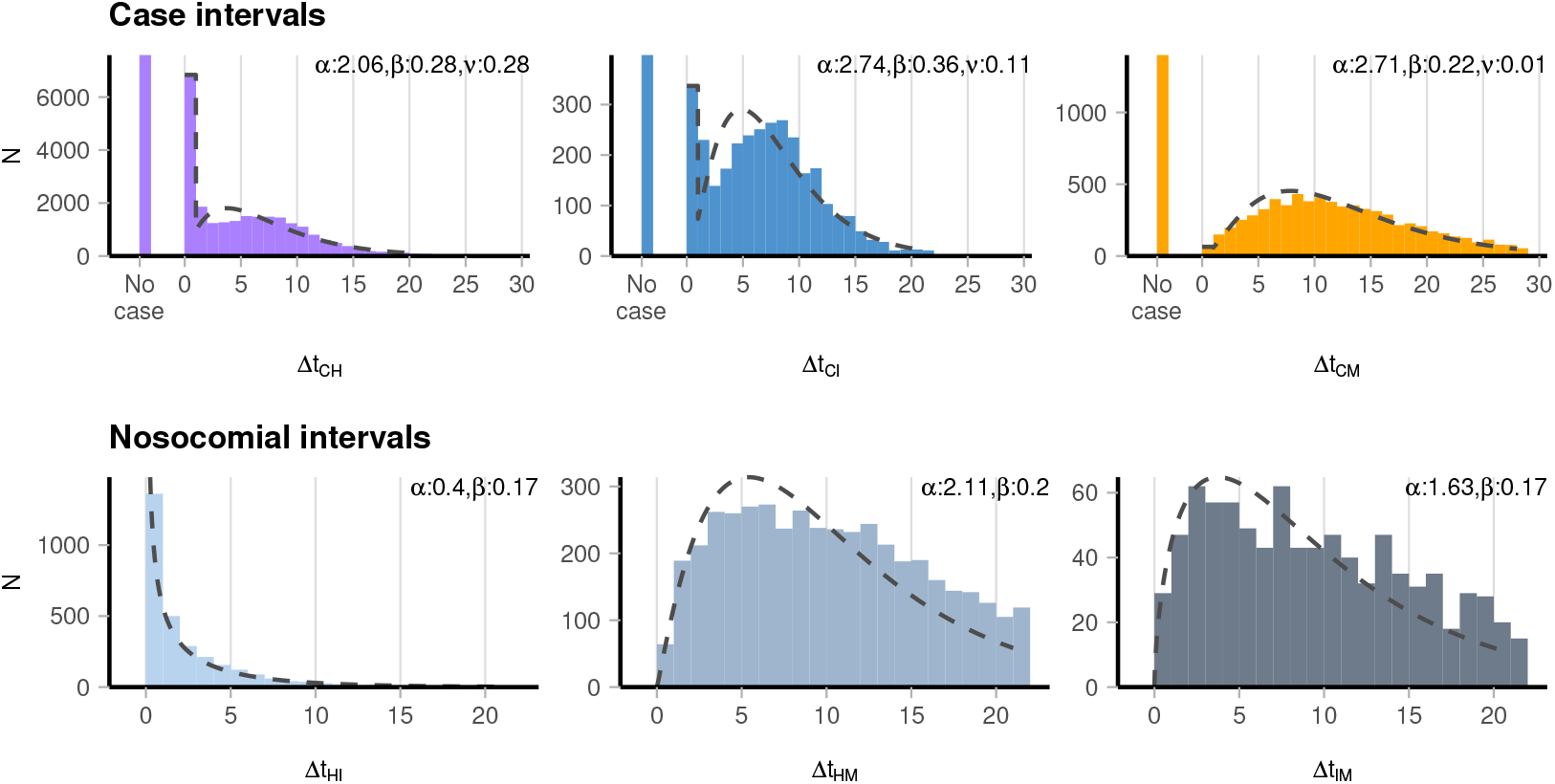
Interval distributions across all ages 20+, comparing time between a (C)ase, (H)ospial admission, (I)CU admission, and (M)ortality. The “no case” entries represent instances where no associated case was found for that severe outcome. Fit parameters are for gamma distributions (with zero inflation for case intervals), detailed in Appendix B.

For outcomes where a corresponding case was found, we first note that for hospital admissions and ICU admissions, a significant proportion (28% and 11% respectively) of these were dated the same day as the case. This proportion remains broadly consistent throughout the period studied (Fig. D.4(b)). *Excluding* these same-day events, the mean case-to-hospitalisation interval (Δ*t*_CH_) was 6.9 days (with 90% of all admissions within the range [1, 15] days). Similarly, the mean case-to-ICU admission interval was 7.3 days [1, 15] and the mean case-to-mortality interval was 11.9 days [2, 24]. 24% of hospital admissions (7,580/31,724), 11% of ICU admissions (398/3,514) and 17% of mortalities (1,399/8,306) had no associated case. These “no-case” entries likely include instances where the individual age/sex/DZ were inconsistent across the data, as well as instances where COVID-19 was identified as a cause, but without a positive test being reported on or before admission.

The same distributions are presented by age, sex, and deprivation of an individual’s residing DZ in Fig. C.1, and Tbls. 1, 2. These reveal that case intervals shorten in older age groups. Focusing on case-to-hospitalisation intervals, 39% (3,814/9,743) of hospital admissions amongst those aged 70+ were on the same day as testing, falling to 16% (936/5,866) for those aged 20–49.

**Table 1:** Case intervals. *Total* : Number of that outcome found in the eDRIS data. *Total linked* : Number of that outcome found with an associated linked case. *Same-day events*: Number of linked events with where the case was reported with the same date. *Median/Mean interval* : Of the linked events found, the median/mean time interval between case and outcome (same-day events excluded). *[5%, 95%]* : Bounding the central 90% of intervals (same-day events excluded).

|  |  | Total | Total linked | Same-day events | Median (d) | Mean (d) | [5%, 95%] |
| --- | --- | --- | --- | --- | --- | --- | --- |
| $\Delta t_{CH}$ | 20-49 | 6895 | 5866 (85%) | 936 (16%) | 7 | 7.4 | [1, 15] |
|  | 50-59 | 5042 | 4243 (84%) | 820 (19%) | 7 | 7.2 | [1, 15] |
|  | 60-69 | 5358 | 4292 (80%) | 1267 (30%) | 6 | 6.8 | [1, 15] |
|  | 70+ | 14429 | 9743 (68%) | 3814 (39%) | 6 | 6.5 | [1, 16] |
|  | Total | 31724 | 24144 (76%) | 6837 (28%) | 6 | 6.9 | [1, 15] |
| $\Delta t_{CI}$ | 20-49 | 868 | 748 (86%) | 72 (10%) | 7 | 7.7 | [1, 15] |
|  | 50-59 | 918 | 817 (89%) | 85 (10%) | 7 | 7.6 | [1, 14] |
|  | 60-69 | 990 | 909 (92%) | 103 (11%) | 7 | 7.0 | [1, 15] |
|  | 70+ | 738 | 642 (87%) | 77 (12%) | 6 | 6.8 | [1, 15] |
|  | Total | 3514 | 3116 (89%) | 337 (11%) | 7 | 7.3 | [1, 15] |
| $\Delta t_{CM}$ | 20-49 | 234 | 179 (76%) | <10 (2%) | 12 | 12.6 | [2, 25] |
|  | 50-59 | 513 | 426 (83%) | <10 (1%) | 11 | 12.3 | [2, 26] |
|  | 60-69 | 1106 | 895 (81%) | <10 (1%) | 12 | 12.6 | [2, 25] |
|  | 70+ | 6453 | 5407 (84%) | 48 (1%) | 11 | 11.8 | [3, 24] |
|  | Total | 8306 | 6907 (83%) | 64 (1%) | 11 | 11.9 | [2, 24] |
|  |  | Total | Total linked | Same-day events | Median (d) | Mean (d) | [5%, 95%] |
| $\Delta t_{CH}$ | Female | 15498 | 11878 (77%) | 3237 (27%) | 6 | 6.9 | [1, 15] |
|  | Male | 16226 | 12266 (76%) | 3600 (29%) | 6 | 6.9 | [1, 15] |
|  | Total | 31724 | 24144 (76%) | 6837 (28%) | 6 | 6.9 | [1, 15] |
| $\Delta t_{CI}$ | Female | 1330 | 1202 (90%) | 116 (10%) | 7 | 7.3 | [1, 15] |
|  | Male | 2184 | 1914 (88%) | 221 (12%) | 7 | 7.3 | [1, 15] |
|  | Total | 3514 | 3116 (89%) | 337 (11%) | 7 | 7.3 | [1, 15] |
| $\Delta t_{CM}$ | Female | 3890 | 3246 (83%) | 26 (1%) | 11 | 11.9 | [2, 24] |
|  | Male | 4416 | 3661 (83%) | 38 (1%) | 11 | 12.0 | [2, 24] |
|  | Total | 8306 | 6907 (83%) | 64 (1%) | 11 | 11.9 | [2, 24] |
|  |  | Total | Total linked | Same-day events | Median (d) | Mean (d) | [5%, 95%] |
| $\Delta t_{CH}$ | 1 (High) | 14952 | 11560 (77%) | 3484 (30%) | 6 | 6.8 | [1, 15] |
|  | 2 (Medium) | 9416 | 7145 (76%) | 1979 (28%) | 6 | 7.0 | [1, 15] |
|  | 3 (Low) | 7356 | 5439 (74%) | 1374 (25%) | 7 | 7.2 | [1, 16] |
|  | Total | 31724 | 24144 (76%) | 6837 (28%) | 6 | 6.9 | [1, 15] |
| $\Delta t_{CI}$ | 1 (High) | 1719 | 1515 (88%) | 187 (12%) | 7 | 7.1 | [1, 15] |
|  | 2 (Medium) | 1051 | 938 (89%) | 89 (9%) | 7 | 7.4 | [1, 15] |
|  | 3 (Low) | 744 | 663 (89%) | 61 (9%) | 7 | 7.6 | [1, 15] |
|  | Total | 3514 | 3116 (89%) | 337 (11%) | 7 | 7.3 | [1, 15] |
| $\Delta t_{CM}$ | 1 (High) | 3773 | 3120 (83%) | 27 (1%) | 11 | 11.8 | [2, 24] |
|  | 2 (Medium) | 2522 | 2113 (84%) | 23 (1%) | 11 | 12.0 | [2, 24] |
|  | 3 (Low) | 2011 | 1674 (83%) | 14 (1%) | 11 | 12.2 | [3, 25] |
|  | Total | 8306 | 6907 (83%) | 64 (1%) | 11 | 11.9 | [2, 24] |

**Table 2:**
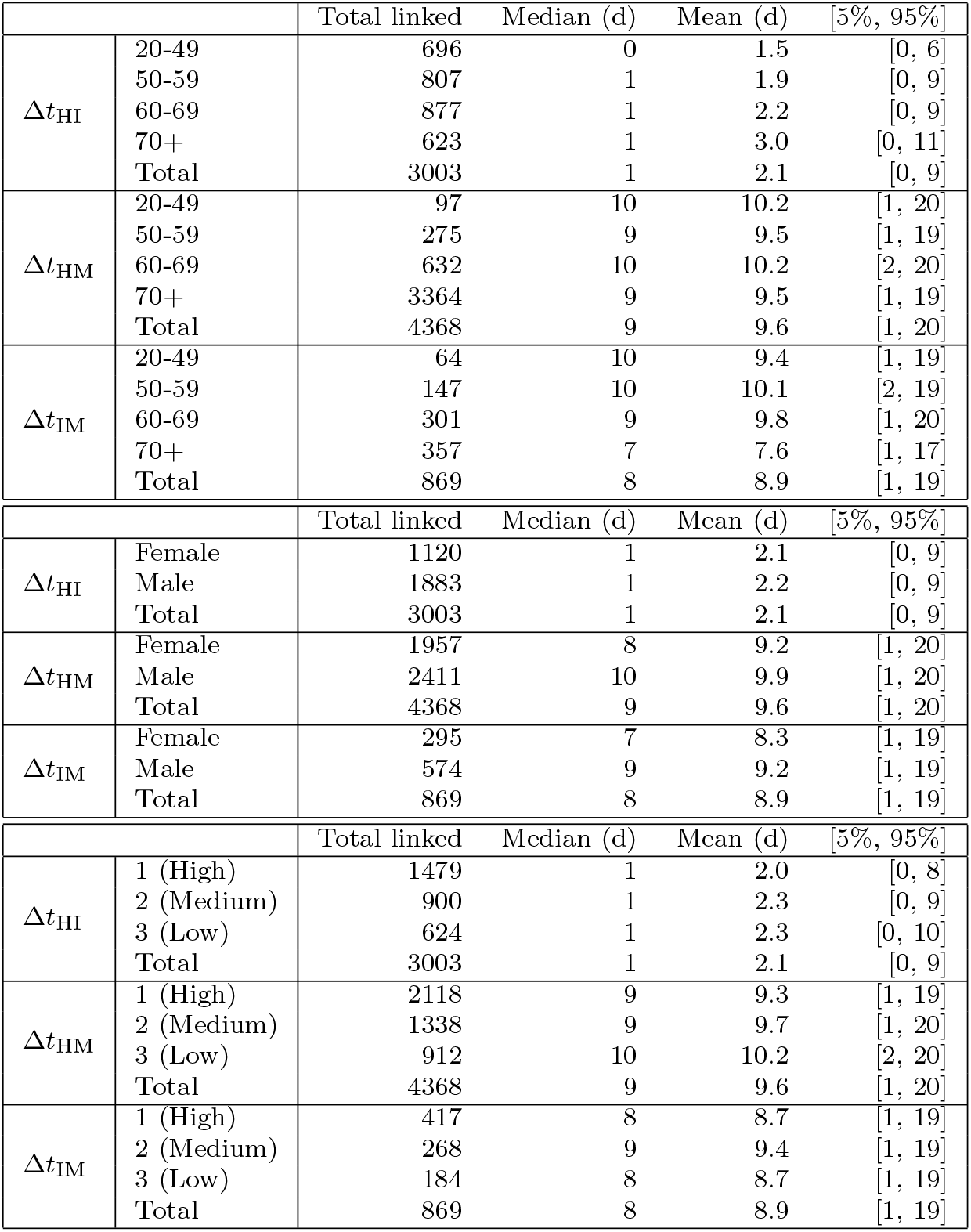
Nosocomial intervals. *Total linked* : Number of that outcome found with an associated prior event in the period studied. *Median/Mean interval* : Of the linked events found, the median/mean time interval between source and outcome. *[5%, 95%]* : Bounding the central 90% of intervals.

There is a smaller difference considering the level of local deprivation, with the proportion of same-day admissions increasing from 25% (1,374/5,439) in the least deprived third of DZs, to 30% (3,484/11,560) in the most deprived third. By sex, 27% (3,237/11,878) of admissions amongst women were on the same day, compared to 29% (3,600/12,266) for men.

Tbl. 2 summarises nosocomial intervals, relating to hospital time of stay. The overall mean interval from hospitalisation to ICU admission Δ*t*_HI_ was 2.1 days [0, 9], with younger age groups having shorter intervals. The mean hospitalisation-to-mortality interval Δ*t*_HM_ was 9.6 days [1, 20]. For those that died after admission to an ICU, the mean interval Δ*t*_IM_ was 8.9 days [1, 19].

### 3.2 Variability in outcomes across different timeframes

During the period studied between September 2020 and January 2022, the epidemic in Scotland evolved in several characteristic ways. First, the dominant coronavirus variant switched three times (the initial *wild* type, followed by the *Alpha* variant introduced around November 2020, followed by the *Delta* variant introduced around May 2021, finally being replaced by the *Omicron* (BA.1) lineage, introduced around November 2021). Additionally, Scotland’s COVID-19 vaccination programme began in December 2020, and by January 1 2022 over 11 million doses had been administered to a population of 5.5 million, with high uptake across all age groups [18]. This was in addition to the introduction of novel antiviral treatments, available to the most vulnerable [19]. The corresponding change in *intervals* throughout the period are then presented in Fig. 2. Case intervals remain broadly consistent, as do the proportion of “same-day” admissions (Fig. D.4). Δ*t*_IM_ falls slightly.

**Figure 2:**
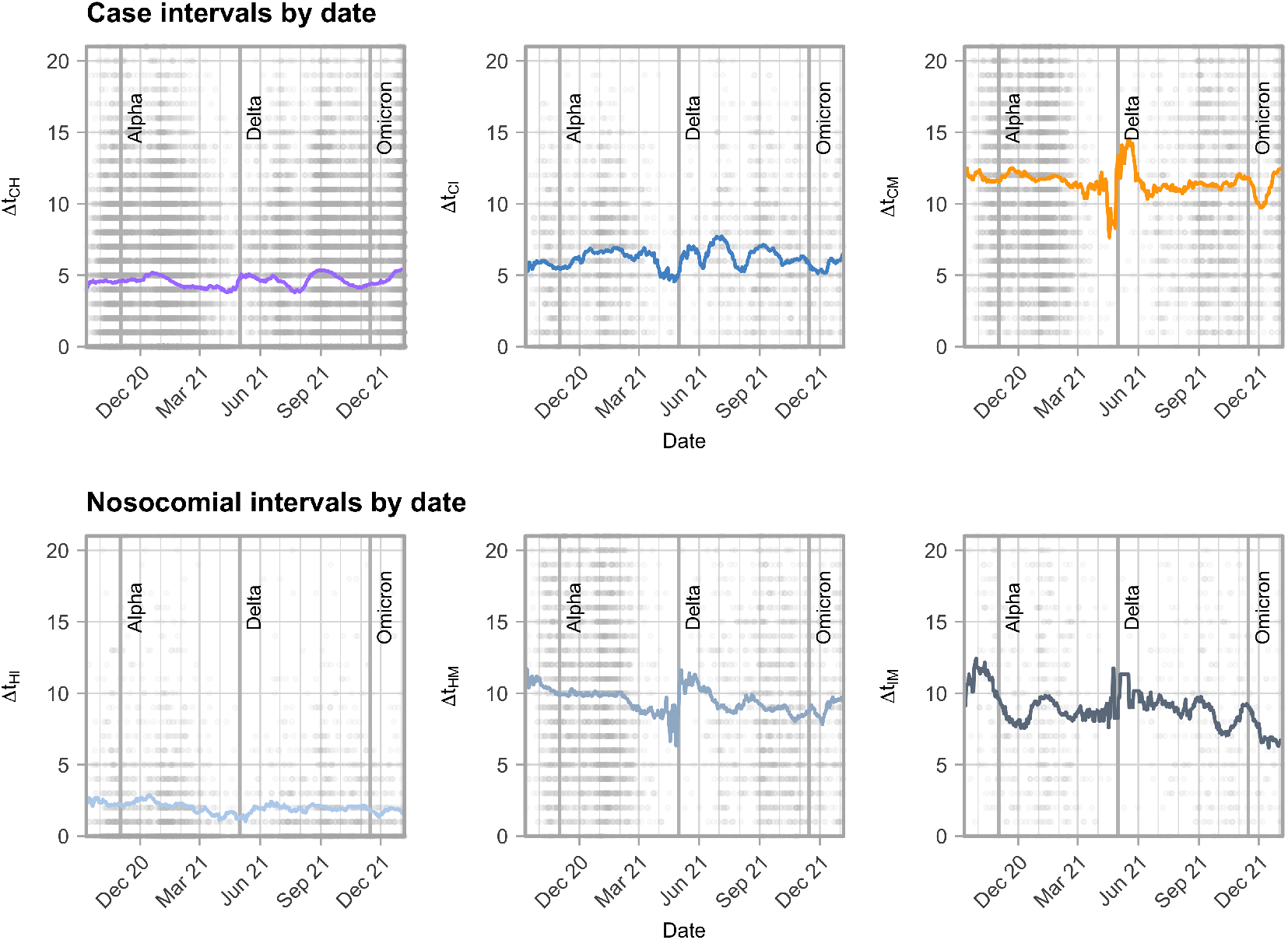
Time evolution of the interval distribution, with points indicating individual instances (taking date of the first outcome, e.g., a case-to-hospitalisation interval is plotted on the date of the case), and the line representing the rolling mean of intervals in a 28-day window centered around that date. The approximate times of introduction of the Alpha, Delta and Omicron variants into Scotland (November 1 2020, May 1 2021 and November 15 2021 respectively) are marked.

### 3.3 Estimated hospitalisation-to-discharge interval

The analysis so far considers explicit intervals between across serial outcomes of increasing severity. The data do not specify date of recovery or discharge, for those admitted to hospital with COVID-19 but later discharged (and assumed to recover). Using a standard approximate Bayesian computation (ABC) algorithm, we estimate a distribution *P* (Δ*t*_HD_) for the nosocomial *hospitalisation-to-discharge* intervals Δ*t*_HD_ for admissions that do not have an associated death, by comparing to the trajectory of hospital *occupancy*, for those with confirmed COVID-19 [17].

Our methodology is detailed in Appendix D. Briefly, we use the eDRIS data to infer admissions of patients that specifically go on to survive (i.e. admissions without an associated mortality) (Fig. D.3(e)), and public occupancy data to infer the occupancy of those patients specifically (Fig. D.3(f)). We then fit a distribution of hospitalisation-to-discharge intervals that best reproduce this occupancy. To account for changes in the epidemic during the period studied, we divide the fit into two periods, with data up to April 30 2021 preceding the first major wave of the *Delta* variant termed the “pre-Delta” period, and data from May 1 2021 onwards the “post-Delta” period.

The posterior distributions (for a zero-inflated exponential distribution with rate *β* and inflation *ν*) are given in Fig. D.2. In the “pre-Delta” period, the mean interval (across all accepted *P* (Δ*t*_HD_)) was E(Δ*t*_HD_) ∼ 13.2 *±* 0.8 days, falling to E(Δ*t*_HD_) ∼ 9.5 *±* 0.5 days in the “post-Delta” period.

Figures D.3(g–h) also show a consistently high proportion of hospital burden (both admissions and occupancy) was from individuals that were eventually discharged (and assumed survived). This proportion is a distinct metric from hospitalisation survival *rates*, as the occupation proportion may also change if the mean length of hospitalisation stay by outcome changed substantially.

We note that in this analysis, hospital admissions obtained from the eDRIS data are marginally lower than the public PHS data across the period. As we discuss further in Appendix D, this may lead to an overestimation of the intervals of each stay.

## 4 Discussion

The COVID-19 pandemic has seen an unprecedented level of consistent, voluntary, recorded testing, in effort to detect and control the spread of infection. This volume of testing, in combination with precise data routinely collected on hospitalisation and mortality, allows us to analyse the natural history of COVID-19 disease at the level of the individual. The monitoring of other infectious diseases in comparison is usually far more rudimentary; the endemic prevalence of influenza in the UK for example is regularly estimated by the volume of phone calls to GPs citing influenza-like symptoms [20]. With the volume of recorded COVID-19 testing since falling to a fraction of that in the period studied here, this period is a unique window for studying intervals of an infectious disease at such detail.

Our work here uses COVID-19 outcomes data in Scotland to draw empirical distributions of intervals between cases and subsequent severe outcomes, as well as intervals between those severe outcomes (e.g., the time between hospital admission and death). The case-to-hospitalisation intervals in particular (Fig. 1) inform, given a surge in COVID-19 cases, how delayed and drawn out a corresponding trajectory in hospital admissions may be. This is important for estimating the timing and size of peak demand on healthcare services (and whether that could exceed capacity), given a surge in cases.

These intervals have a clear age structure (Fig. C.1, Tbl.1). Notably, this variation is largely seen in the proportion of “same-day” events; 16% of all age 20–49 admissions were on the same day compared to 39% for ages 70+, whereas the mean interval for those not admitted on the same day only fell slightly, from 7.4 days to 6.5 days. This larger same-day variation is also seen with respect to deprivation. This suggests a difference in risk profiles between those testing prior to admission (where severe symptoms develop later), and those testing on the same day or on admission. A likely influencing factor is admissions from those admitted to hospital for a non-COVID-19 reason, but tested positive on a routine test on admission. Prior to the introduction of the Omicron variant the proportion of COVID-19 admissions being “with” COVID-19 remained at approximately 25% (Fig. D.4(a)).

As well as age, deprivation and sex have been highlighted as risk factors for poor COVID-19 outcomes, with higher rates associated with men [21, 22, 23], and individuals living in more deprived communities [2, 24, 25]. Our analyses confirm this in the overall number of severe outcomes per group, and reveal modest variation in their intervals (Tbl. 1), with a slightly higher proportion of same-day admissions from both men, and individuals living in more deprived communities.

Compared to cases (dated to when a test is *taken*), the delay from *infection* or *onset of symptoms* to these severe outcomes will be longer, taking into account the incubation period of the disease (estimated to have a median of order five days [26, 27]), and potential delay between onset of symptoms and taking a test. In light of this, our results are then broadly consistent with intervals presented by a UK study [15], which infers the mean interval from infection to hospitalisation prior to January 2021 to have been of order 8–10 days (as compared with our mean case-to-hospitalisation interval of 6.9 days), and time from infection to mortality to have been of order 9–16 days (compared to our mean case-to-mortality interval of 11.9 days). A more recent work in Reference [16] studies intervals in South Korea over a similar period to that studied here, and finds a longer mean interval from symptom onset to death of 20.1 days, and from symptom *reporting* to death of 16.7 days. The measured interval from cases or onset to more severe outcomes will have several influencing factors. These include but are not limited to the frequency of testing (in turn the mean time after infection that a test is taken), the properties of the virus and differences across different variants and, as highlighted here, the structure of the population with respect to age. It is therefore reasonable to expect variation in absolute values, but trends such as a shortening in intervals with increasing age appear to be consistent across studies [15, 16].

While the *nosocomial* intervals — relating to time of stay in hospital — are routinely collected, they are not often available to researchers at such detail, with the data used here an exception owing to the need for modelling support in the acute stage of this pandemic. We see variation in these intervals through the period studied. For those admitted to hospital that go on to die, the interval from admission to death shortens over the period (Fig. 2). This is somewhat counter-intuitive given improving rates of survival after admission (Fig. D.3(g–h)), but may be due to individuals who recover and are discharged, that may have died if infected at an earlier stage of the pandemic (where treatment of severe COVID-19 disease was less well understood). If one then assumes that these “removed’ individuals are generally healthier and would have stayed alive in hospital for longer, the removal of these individuals would reduce the mean interval overall.

The programme of free community testing in Scotland has allowed the spread of the virus to be tracked at remarkable spatio-temporal resolution [28, 29, 30, 2, 31, 32, 33]. Despite prevalence remaining high throughout 2022 [34], Scotland has since entered a phase where testing is no longer mandatory, nor generally free of charge. The proportion of COVID-19 infections being identified is now very low. The absence of such testing data may result in future outbreak modelling relying on the trajectories of severe outcomes only, or more basic estimates of incidence. Combined with a reasonable understanding of the incubation period and potential case ascertainment, our interval distributions may help infer routes of transmission and patterns of infection.

## Data Availability

The outcomes data utilised in this work are not publicly available. They are provided to the authors for academic research by Public Health Scotland's electronic Data Research and Innovation Service, under a data sharing agreement (Spatial and Network Analysis of SARS-CoV-2 Sequences to Inform COVID-19 Control in Scotland). Deprivation data are obtained from the 2020 Scottish Index of Multiple Deprivation, which is publicly available (https://simd.scot). Nationwide-level occupancy of patients with COVID-19 are obtained from Public Health Scotland, and is also publicly available (https://www.gov.scot/
publications/coronavirus-covid-19-trends-in-daily-data/).

https://www.gov.scot/publications/coronavirus-covid-19-trends-in-daily-data/

## 5 Acknowledgements

We thank eDRIS for the provision of COVID-19 testing and severe outcomes data.

## 6 Author contributions

R.R.K. conceived the project. A.J.W. wrote the underlying code and performed the analysis, and with R.R.K. wrote the manuscript.

## 8 Code availability

Analysis code is available at https://git.ecdf.ed.ac.uk/awood310/covid-19-outcomes-intervals-analysis.

## 9 Data availability

The outcomes data utilised in this work are not publicly available. They are provided to the authors for academic research by Public Health Scotland’s electronic Data Research and Innovation Service, under a data sharing agreement (*Spatial and Network Analysis of SARS-CoV-2 Sequences to Inform COVID-19 Control in Scotland*). Deprivation data are obtained from the 2020 *Scottish Index of Multiple Deprivation*, which is publicly available [35]. Nationwide-level occupancy of patients with COVID-19 are obtained from Public Health Scotland, and is also publicly available [17].

## A Associating outcomes

We define an interval Δ*t*_AB_ ≥ 0 as the time difference between two different COVID-19 outcomes *A* and *B*, given as a whole number of days. We do not differentiate by any intermediate outcomes; for example, the case-to-mortality intervals includes both patients that were and were not admitted to an ICU.

We link different events with one another. For example, consider a hospitalisation entry *H*, for which we are attempting to associate a case *C*. To do this we:

1. Search for cases {*C*} from the eDRIS test data, where the DZ, age range and sex matches with *H*, and occurred on the same day as, or up to 28 days before *H*.
2. If at least one one possible matching case is found, take the interval Δ*t*_CH_ as the time difference between *H* and and the median date of the candidate cases {*C*}. Otherwise, label the hospitalisation entry *H* as *unlinked*.

We associate outcomes up to 21 days apart, with the exception of case-to-mortality intervals, where we search over 28 days. For case intervals, *unlinked* instances are reports of more severe outcomes, but without an associated prior case reported. For nosocomial intervals, unlinked outcomes are common (such as a mortality without an ICU admission), and are not counted.

Finally, in the data we omit events with incomplete age/sex/DZ entries (as we use these to associate different outcomes), as well as repeat admissions by the same individual within a window of 60 days, taking only the first admission.

## B Distribution fits

For fitting the empirical distributions from Figures 1,C.1, we choose two-parameter *gamma* distributions *P* (Δ*t*):

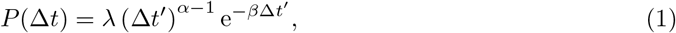

for 0 ≤ Δ*t* ≤ 21 days (28 days for Δ*t*_CM_) and zero outside this range, with *α* determining the characteristic shape of the distribution for smaller Δ*t*, and *β* determining the rate of exponential decay as Δ*t* increases, and *λ* a normalising constant fixing 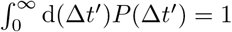.

To account for instances where the case and a more severe outcome are on the same day (Δ*t* = 0), case intervals are fit across Δ*t* ≥ 1 only, with a *zero-inflation ν* fit separately, to reflect the proportion of all “same-day” events:

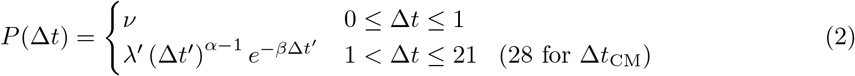

where *λ*^*′*^ here fixes 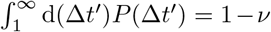. These fit values are given alongside the distributions in Figures 1, C.1.

## C Intervals by age, sex and local deprivation

Statistics on intervals are given in Tbl. 1 for case intervals, and Tbl. 2 for nosocomial intervals. These are specified by age, sex and the relative deprivation of residing DZ (equally dividing the DZs into three groups, based on the overall rank in the *Scottish Index of Multiple Deprivation* [35]). We finally plot the intervals by age group in Fig. C.1.

## D Estimation of hospitalisation-to-discharge intervals

In this section we detail the method to estimate a distribution for the interval between hospitalisation and discharge, for patients presumed to not die in hospital. This is a much broader estimate across the whole population, as we do not have explicit times between admission and discharge. We instead rely on public, national-level occupancy data, provided by PHS [17].

For those admitted with COVID-19 that go on to die, we first use the eDRIS data (and associated intervals between hospital admission and death) to derive a partial occupancy timeseries (Fig. D.3(d)). The difference between this occupancy and the *overall* PHS occupancy is then taken as the occupancy of admitted individuals that are discharged (Fig. D.3(f)). Finally, knowing the admission dates of patients that go on to survive (i.e., do not have an associated death) from the eDRIS data, we estimate the hospitalisation-to-discharge interval distribution, and thus how much surviving individuals on average contribute to the hospitalisation occupancy burden (Fig: D.3(h)).

Formally, the trajectory of hospital admissions *A*(*t*) includes those that go on to be discharged (and we assume recover) *A*_D_(*t*), and those that die in hospital *A*_M_(*t*):

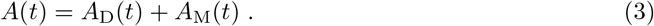

Similarly, the trajectory of COVID-19 hospital occupancy *O*(*t*) includes the occupancy of those that go on to recover and be discharged *O*_D_(*t*) and those that go on to die *O*_M_(*t*):

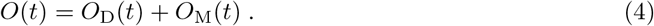

To estimate the discrete distribution *P* (Δ*t*_HD_) for the (H)ospital admission-to-(D)ischarge interval, we first rewrite the occupancy of those eventually discharged

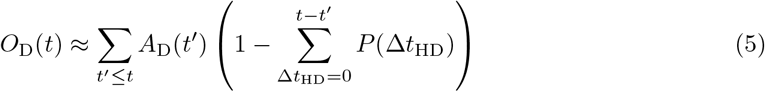

writing *O*_D_(*t*) as a *convolution* of the admissions trajectory *A*_D_(*t*^*′*^), and the proportion of individuals remaining in hospital after interval *t* − *t*^*′*^. (1 − *P* (0)) is then the proportion of patients that are in hospital for at least one full day. Knowing *O*_D_(*t*) and *A*_D_(*t*), what remains is to fit an appropriate *P* (Δ*t*_HD_).

We assume *P* (Δ*t*_HD_) follows a zero-inflated exponential distribution, with the zero inflation accounting for individuals admitted, but discharged without an overnight stay. We use a standard Approximate Bayesian Computation (ABC) algorithm in a two-parameter space (exponential decay rate *β*, and zero-inflation *ν*), and take two different fits for the periods September 10 2020 – April 30 2021, and May 1 2021 – January 6 2022.

The prior (for *ν*: *U* (0, 1) and for *β*: *U* (0.05, 1)), allows for any zero-inflation, and restricts the mean occupancy of those that stay at least one night between 1.1 and 20 days. We build a posterior by accepting parameters that generate modelled occupancy trajectories *Õ*_D_(*t*) that best fit *O*_D_(*t*). We sample 10^6^ different pairs of parameters, from which we take the 1, 000 that produce via Eq. (5) the timeseries 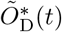 that best fits *O*_D_(*t*), minimising the sum of absolute residuals

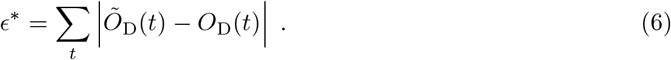

Between September 10 2021 and January 6 2022, PHS reported a total of 38,480 admissions, whereas 32,718 were derived from the eDRIS data in the same period. With fewer admissions (Fig. D.3(a)), we may then over-estimate the mean interval of each stay. There is also a clear time dependence; in Fig. D.3, the admission peak of 241 on January 11 2021 preceded an occupation peak of 2,053, 11 days later. However, a second admissions peak of 202 on September 7 2021 preceded a much lower occupation peak of 1,107, 14 days later, suggesting those admitted were staying for less time. We have then separately fit two periods, but Fig. D.3(f) suggests further variation within these periods. From December 2021 onward, hospital admissions are most likely attributable to infection with the *Omicron* variant, where material changes in admissions trends would be expected as compared to previous variants, owing to a lower observed severity [36].

**Figure C.1:**
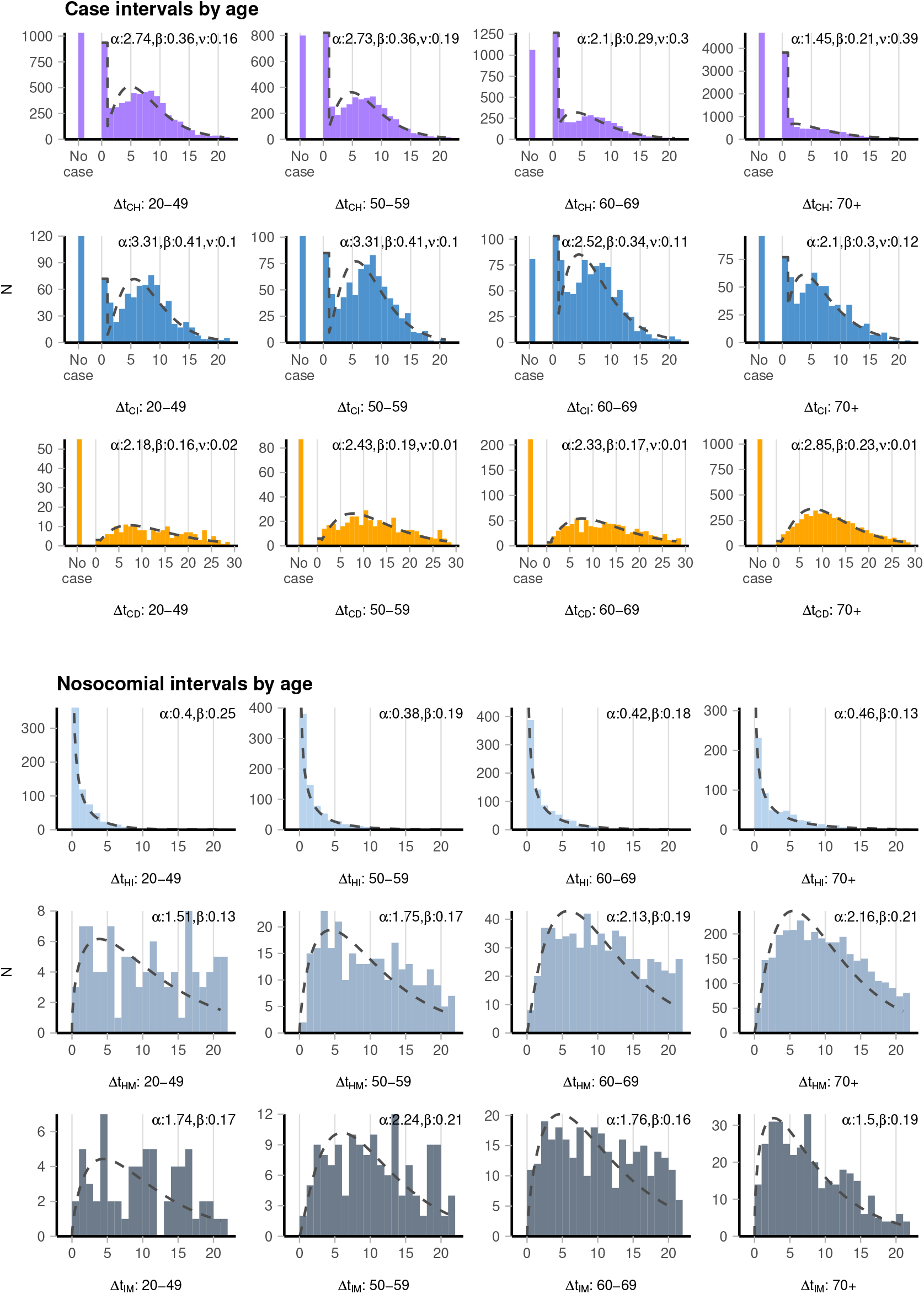
Interval distributions, separated by age group. A “no-case” entry is logged in the case intervals when a severe COVID-19 outcome (such as a hospitalisation) is identified, but has no associated case. *α, β*, are fit values for the gamma distribution rate, shape and *ν* is the zero inflation parameter (if applicable).

**Figure D.2:**
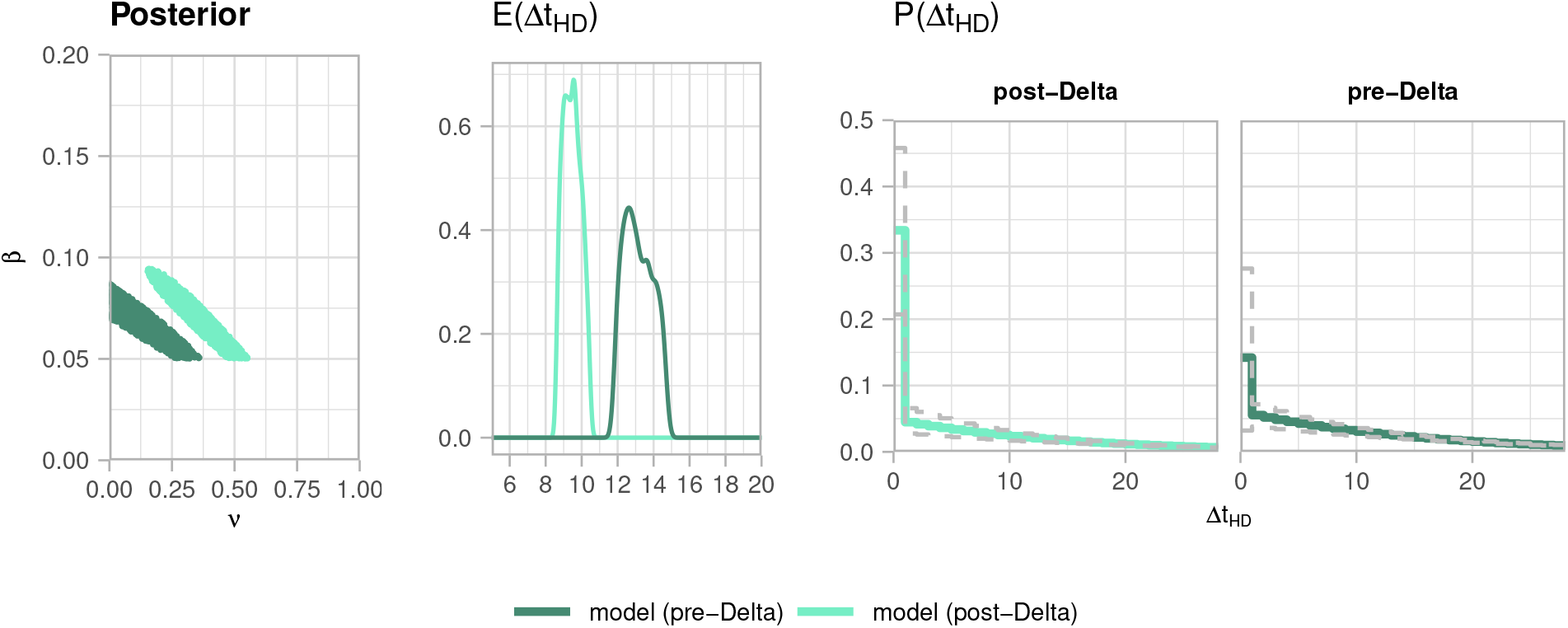
Posterior distributions, mean intervals and distributions for hospitalisation-to-discharge intervals Δ*t*_HD_, across the “pre-Delta” (10 September 2020 – 30 April 2021) period, and “post-Delta” period (1 May 2021 – 6 January 2022).

**Figure D.3:**
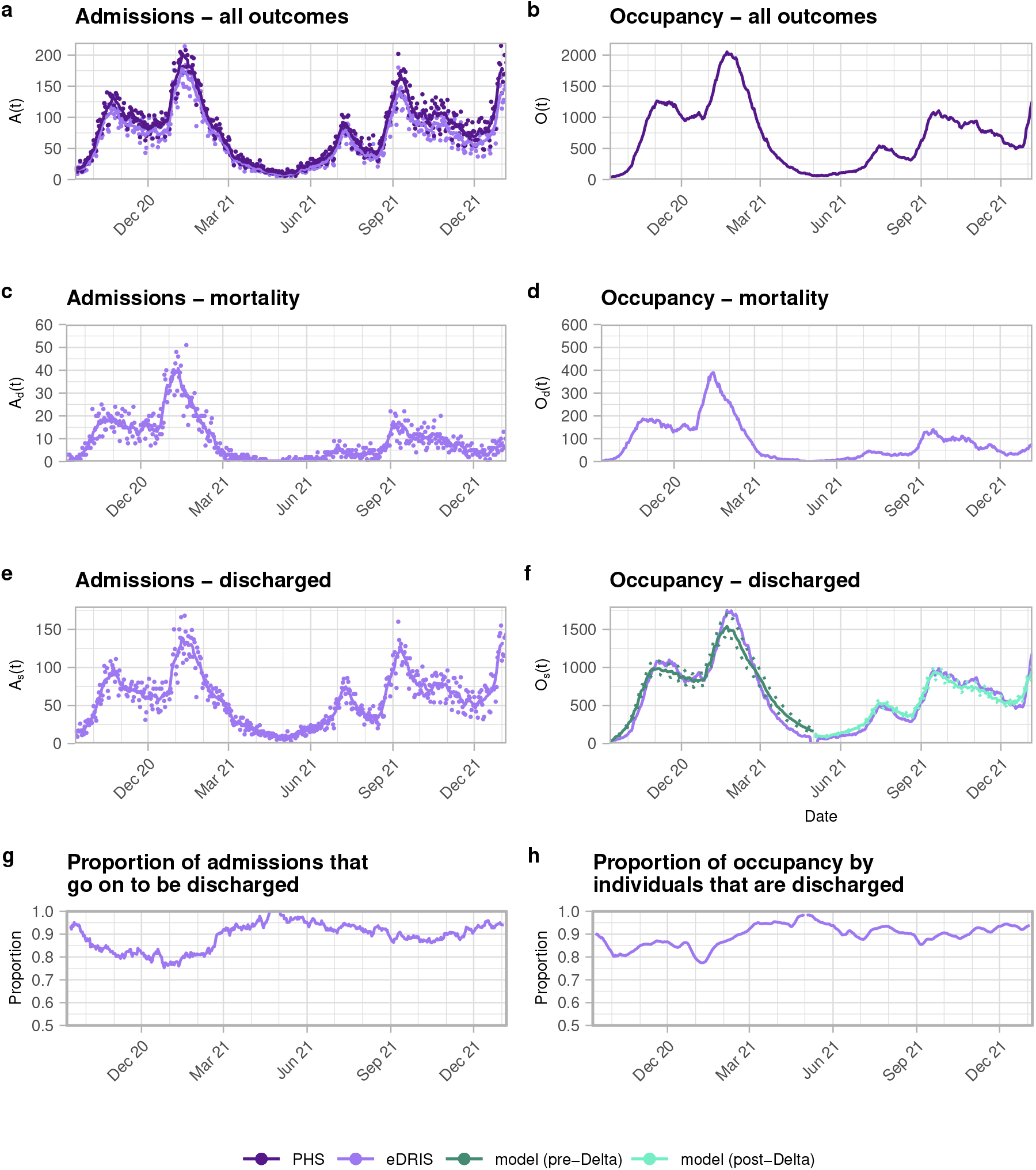
Trajectories of hospital admissions, and occupancy, of COVID-19 patients in Scotland. Overall admissions (a) and occupancy (b) are from Public Health Scotland (purple). PHS-published admissions are generally higher than those derived from the eDRIS data stream (magenta). We then infer which of those admissions were from patients that eventually died (c), and the hospital occupancy of those individuals (d). The remaining admissions (e) and occupancy (f) are then taken to be from patients that go on to be discharged (and we assume survive). Finally the proportion of admissions (g) and occupancy (h) by patient outcome.

**Figure D.4:**
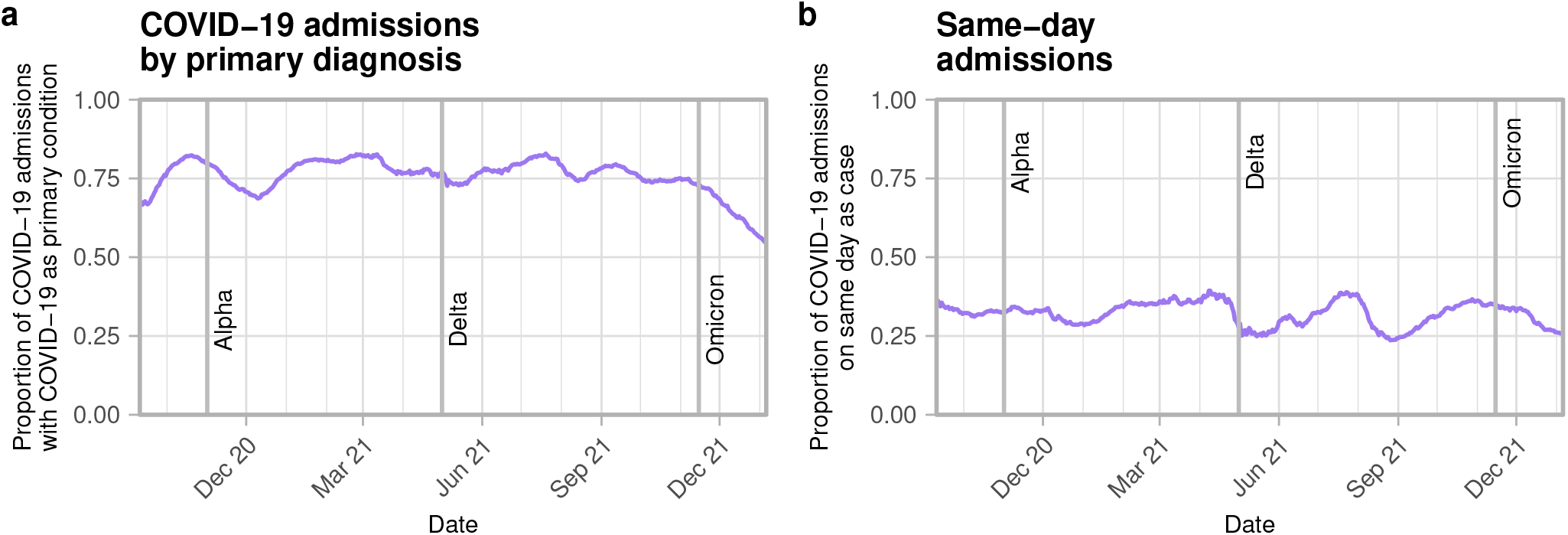
28-day rolling mean of proportion of COVID-19 hospital admissions by (a) whether COVID-19 was the primary reason for admission, (b) whether the admission was on the same day as the related COVID-19 case (for admissions with an associated case). The approximate times of introduction of the Alpha, Delta and Omicron variants into Scotland (November 1 2020, May 1 2021 and November 15 2021 respectively) are marked.

